# The impact of asthma on mental health & wellbeing during COVID-19 lockdown

**DOI:** 10.1101/2020.09.10.20190793

**Authors:** Daniel H Higbee, George Nava, Alex S F Kwong, James W Dodd, Raquel Granell

## Abstract

**Introduction:** The global SARS CoV2 pandemic resulted in social isolation measures with unintended negative impacts, particularly on mental health. We hypothesised that people with asthma are likely to be more vulnerable to worse mental health during lockdown.

**Methods:** We examined COVID-19 surveys (completed April/May 2020), nested within two generations of the Avon Longitudinal Study of Parents and Children (ALSPAC): index-generation ALSPAC-G1 (n= 2942, mean age=28) and the parent’s generation ALSPAC-G0 (n=3737, mean age=59). We used Poisson and logistic regression models to estimate the effect of asthma on wellbeing, anxiety and depression, and factors related to COVID-19 and lockdown. Models were adjusted for validated pre-pandemic measures of mental health and socio-economic factors.

**Results:** Asthma was associated with a 13% increase in depression score in ALSPAC-G1 (p=0.005) and 15% increase in ALSPAC-G0 (p=0.05) compared to participants without asthma, anxiety scores increased by 14% in ALSPAC-G1 (p=0.005) and by 16% in ALSPAC-G0 (p=0.02). Asthma was associated with a similar increase of anxiety and depression scores during COVID-19 in both generations (Z test p values >0.80).

**Discussion:** People with asthma have worse mental health & wellbeing during lockdown compared to people without asthma. Although the effect of asthma on mental health is of similar magnitude between the generations, younger participants with asthma declined to lower levels of mental health despite reporting less symptoms, COVID-19 infection and self-isolation. This has important implications given repeated lockdowns. Young people with asthma should be closely monitored and supported to mitigate the impact of lockdown on their mental health.

**Key Messages:** 

**What is the question?:** What is the impact of asthma on mental health & wellbeing during COVID-19 pandemic?

**What is the bottom line?:** People living with asthma report worse wellbeing, anxiety and depression in lockdown compared to those without asthma, the effect is not entirely explained by pre-existing mental health problems, physical symptoms or COVID-19 infections.

**Why read on?:** Young people living with asthma are more likely to report concerns about susceptibility to COVID 19 and job security. The negative impact of asthma on length of self-isolation, suspected COVID and symptoms appears greater in older people with asthma.

## Introduction

The novel severe acute respiratory syndrome coronavirus 2 (COVID-19) pandemic has resulted in millions of infections and hundreds of thousands of deaths [1]. It has been assumed that people living with pre-existing respiratory diseases would be at higher risk of developing severe illness, despite a lack of available evidence [2]. To mitigate this possible risk, and to reduce transmission, measures to restrict social interaction including social distancing and self-isolation were introduced. Similar measures were used during the Severe Acute Respiratory Syndrome (SARS) epidemics leading to negative mental health impacts even in those not exposed to the virus [3]. Unlike SARS, COVID-19 has caused a high proportion of the global population to endure social isolation measures and financial instability, increasing the potential for psychological harm. Some regions of the UK, and countries around the world, have re-instigated social isolation measures causing an ongoing potential threat to mental health.

Asthma is a common respiratory disease of multi-morbidity affecting ~10% of the UK’s population and is associated with anxiety and depression [4, 5]. The presence of psychological co-morbidity with asthma decreases quality of life, increases healthcare utilisation and contributes to the risk of asthma death [6-8].

Cross sectional studies have shown a high prevalence of anxiety and depression during COVID-19 across ages and countries [9, 10]. Available sample populations thus far have not included longitudinal data (including pre pandemic measures) or detailed clinical history, prior physical or psychological assessment, meaning that conclusions about the effect of COVID-19 on people with asthma have been impossible.

Our objective was to determine if people with asthma experienced worse anxiety, depression and/or wellbeing during COVID-19 compared to people without asthma and to explore the additional impact of the COVID-19 pandemic on associated factors. We hypothesize that physical symptoms such as having a suspected COVID-19 infection or difficulty breathing, as well as social factors such as worry of losing a job, could impact people with asthma more severely than people without asthma. We utilised data from a British longitudinal cohort of children born in 1991 and their parents to compare associations in young adults and middle aged [11]. This cohort has been extensively investigated at regular time points allowing us to compare pre-COVID-19 levels of mental and physical disease to anxiety and depression assessed during lockdown. In April 2020, ALSPAC participants completed a questionnaire covering COVID-19 symptoms, mental health, lifestyle, finances, and family life. This questionnaire was designed to quantify the impact of COVID-19 and learn more about the psychological, social, and economic impact of the government’s lockdown strategy.

We used this COVID-19 questionnaire data along with a wealth of longitudinal pre-pandemic data. If more severe worsening of mental health or wellbeing is evident among people with asthma, it could be used to help identify those in need of additional support. It is also important to understand other factors which may be associated with changes in mental health to determine if they may be modifiable. Results may also help balance public health decisions regarding re-instigation of lockdown measures.

## Methods

### Study Samples

The Avon Longitudinal Study of Parents and Children (ALSPAC) is an ongoing longitudinal population-based study that recruited pregnant women residing in Avon in the south-west of England with expected delivery dates between 1st April 1991 and 31st December 1992 [11, 12]. The cohort consists of 13,761 mothers and their partners (hereafter referred to as ALSPAC-G0), and their 14,901 children (ALSPAC-G1) [13]. Details are available in the **Supplementary material, Appendix 1**.

This study uses data from 3737 ALSPAC-G0 and 2942 ALSPAC-G1 who completed an online questionnaire about the impact and consequences of the COVID-19 pandemic between 9th April and 14th May 2020 [14]. Lockdown was announced in the UK on the 23^rd^ March.

### Self-reported Current Asthma

Current asthma was assessed in the COVID-19 surveys with the question-Participant has asthma yes/no- at mean age 28 years in ALSPAC-G1 and at mean 59 years in ALSPAC-G0. In ALSPAC-G1 the proportion of participants with asthma correlated well with previous reports of current asthma at 23 years (92% overlap); in ALSPAC-G0 the overlap with previous report of asthma was 72% with ‘asthma ever’ at 39 years.

### Pre-COVID-19 vs COVID-19 Mental Health

The measures used in the COVID-19 survey examine symptoms in the preceding 2 weeks, thus represent mental health in lockdown. Depressive symptoms in both samples were measured using the Short Mood and Feelings Questionnaire (SMFQ) [18]. Scores range between 0-26 with higher scores indicting higher depressive symptoms. Anxiety symptoms were measured using the Generalised Anxiety Disorder Assessment (GAD-7) [19]. Scores range between 0-21 with higher scores indicting higher anxiety symptoms. Mental wellbeing was measured using the Warwick-Edinburgh Mental Wellbeing Scale (WEMWBS) [20]. Scores range between 14-70, with higher scores indicating better mental wellbeing.

In ALSPAC-G1 the same scores measured in the COVID-19 survey (mean age=28) were available pre-pandemic: WEMWBS-14 at 24 years, SMFQ-13 score at 26 years and GAD-7 score at 22 years.

In the older ALSPAC-G0 population (mean age=59), mental wellbeing was not previously assessed. We used Edinburgh Postnatal Depression Scale score for depression [15] at 52 years (range 0-30) and State-Trait Anxiety Inventory score for anxiety [16] at 39 years (range 20-80), as pre-pandemic measures for adjustment. See **Appendix 2** for more information.

### Symptoms, Change in Activities and Worries during lockdown

We explored the differences in confirmed or suspected COVID-19, shortness of breath or difficulty in breathing in last week, contact with confirmed or suspected COVID-19 in last 2 weeks, difficulty sleeping in last week, severe fatigue in last week, went to A&E about symptoms last week, sought medical attention last week), change in activities (self-isolation, length of self-isolation, sleep/exercise/alcohol drinking/eating change since lockdown) and worries during lockdown (worried about getting COVID-19/losing their job, in participants with and without asthma.

### Statistical analyses

Analysis was conducted in Stata (version 15) [17].

We reported p-values from Pearson Chi-square test when comparing categorical characteristics and Z-test from logistic regression when comparing continuous characteristics in people with asthma vs. people without asthma.

We used Two-sample Kolmogorov-Smirnov test for equality of distribution functions to compare mental health scores between the younger population with asthma (ALSPAC-G1) vs. the older population with asthma (ALSPAC-G0).

We used Poisson and logistic regression models to estimate the effect of asthma on wellbeing, anxiety and depression, and other factors relating to COVID-19 and lockdown. We did not impute missing data as a different analysis using the same data resources did not find it changed results [18]. Analysis was conducted separately for ALSPAC-G0 and ALSPAC-G1, adjusting for sex, age, smoking/vaping status, overweight and pre-existing mental health. Exponentiated Poisson regression estimates, exp(b), were reported as symptom count ratios (SCRs), known as incident rate ratios. These can be interpreted as a percentage increase/decrease of the outcome score in participants with asthma, compared to participants without asthma given as (SCR-1)×100 [19, 20]. Estimates and corresponding 95% confidence intervals from Poisson and logistic regression models for ALSPAC-G0 and ALSPAC-G1 were displayed together in **Figure 1**, further details in **Supplementary information, Appendix 3**.

**Figure 1.**
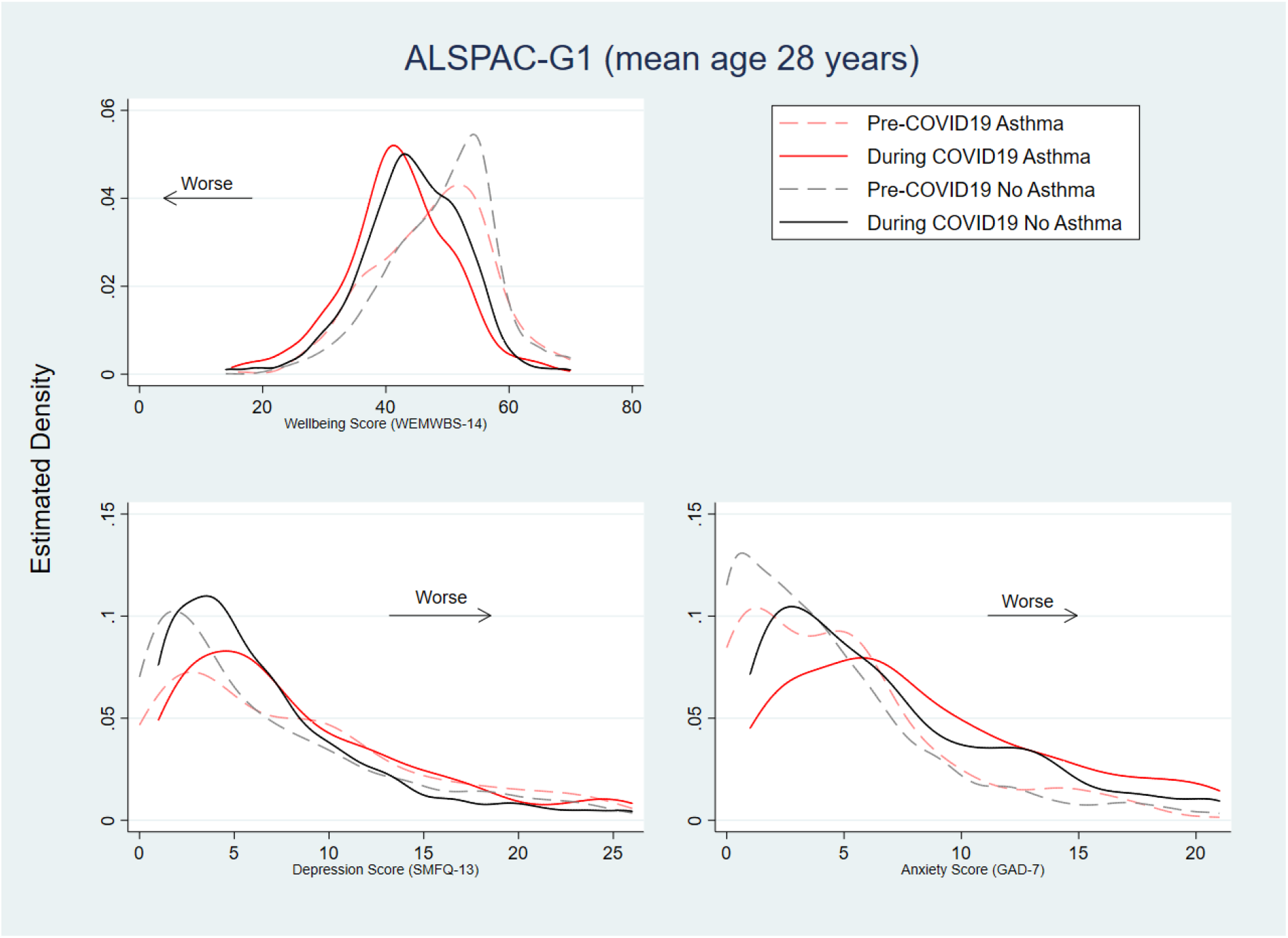
Kernel density estimate plot of mental health scores in ALSPAC-G1 children with and without asthma, pre and during COVID-19. **Figure 1**. shows lower wellbeing and depression scores in those with asthma pre-COVID-19, compared to those without asthma. Anxiety scores were similar. During COVID-19 mental health worsened in both groups, but there was a greater decline to a lower level in those with asthma.

We used 2-sample Z-test to compare the effect of asthma in the younger population (ALSPAC-G1) vs. the older population (ALSPAC-G0).

## Results

### Demographics

410 (13.9%) participants in ALSPAC-G1 (mean age 28) and 400 (10.7%) in ALSPAC-G0 (mean age 59 years) responded yes to having asthma in the COVID-19 survey. Demographics are reported in **Table 1**. Participants with asthma reported being overweight more (28% vs. 19% p=1.4×10^-5^ in ALSPAC-G0 and 39% vs 33% p=0.02 in ALSPAC-G1) and taking regular medications (72% vs. 40% p=1.8×10^-32^ in ALSPAC-G0 and 89% vs. 60% p=1.4×10^-29^ in ALSPAC-G1), compared to participants without asthma. Furthermore, in ALSPAC-G0 we reported more females (79% vs. 72% p=0.01) and keyworkers (38% vs. 31% p=0.01) among people with asthma, compared to those without.

**Table 1.** Demographics in G1-Index children and G0 parents’ cohorts

|  | G1-Index Children |  |  |  |  | G0 Parents |  |  |  |  |
| --- | --- | --- | --- | --- | --- | --- | --- | --- | --- | --- |
|  | No Asthma at 28 years |  | Asthma at 28 years |  |  | No Asthma at 59 years |  | Asthma at 59 years |  |  |
|  | N=2532 (86.1%) |  | N=410 (13.9%) |  |  | N=3337 (89.3%) |  | N=400 (10.7%) |  |  |
|  | Total | Mean (SD) | Total | Mean (SD) | Pvalue† | Total | Mean (SD) | Total | Mean (SD) | Pvalue† |
| Age in years | 2532 | 27.61 (0.54) | 410 | 27.57 (0.54) | 0.18 | 3337 | 58.73 (4.78) | 400 | 57.79 (4.95) | 2.13E-04 |
|  | Demographics at 28 years |  |  |  |  | Demographics at 59 years |  |  |  |  |
|  | Total | N (%) | Total | N (%) | Pvalue† | Total | N (%) | Total | N (%) | Pvalue† |
| Gender (male) | 2530 | 727 (28.7) | 410 | 112 (27.3) | 0.56 | 3337 | 917 (27.5) | 400 | 85 (21.3) | 0.008 |
| Smoking/vaping | 2479 | 494 (19.9) | 403 | 85 (21.1) | 0.59 | 3228 | 297 (9.2) | 378 | 27 (7.1) | 0.19 |
| Overweight | 2517 | 471 (18.7) | 407 | 114 (28) | 1.36E-05 | 3309 | 1079 (32.6) | 399 | 154 (38.6) | 0.016 |
| Diabetes | 2528 | 11 (0.4) | 408 | 5 (1.2) | 0.04 | 3324 | 130 (3.9) | 397 | 21 (5.3) | 0.19 |
| Keyworker | 2393 | 1303 (54.5) | 392 | 215 (54.8) | 0.88 | 3122 | 967 (31) | 375 | 141 (37.6) | 0.01 |
| Taking regular medications | 2530 | 1014 (40.1) | 410 | 293 (71.5) | 1.83E-32 | 3334 | 1997 (59.9) | 400 | 355 (88.8) | 1.42E-29 |

† Pearson Chi-square Test for categorical variables, Z-test from logistic regression for continuous variables

### Pre-COVID-19 vs COVID-19 Mental Health

The younger ALSPAC-G1 participants with asthma reported worse pre-existing wellbeing at 24 years (mean (SD) 47.5 (9.4) vs. 49.0 (8.7) p=0.01) and worse pre-existing depression at 26 years (8.1 (6.6) vs. 6.6 (6.2) p=1.6×10^-4^), compared to participants with no asthma. We observed little or no evidence for worse pre-existing anxiety scores at 22 years (p=0.22). However, in the COVID-19 survey G1 participants with asthma reported worse wellbeing (42.2 (8.8) vs. 44.4 (8.4) p=1.9×10^-6^), more depression (8.3 (6.2) vs. 6.7 (5.3) p=1.6×10^-7^) and more anxiety (8.3 (5.4) vs. 6.9 (5.0) p=1.9×10^-6^), compared to younger people with no asthma (see **Table 2 and Figure 1**). In the older ALSPAC-G0 population, we observed little evidence of people with asthma reporting worse pre-COVID-19 lockdown depression or anxiety (p≥0.06), however they did report worse depression (3.6 (4.4) vs. 2.9 (3.6) p=4.5×10^-4^) and anxiety (4.2 (4.9) vs. 3.4 (4.0) p=2.3×10^-4^) at the lockdown COVID-19 survey, compared to older people with no asthma (see **Table 2**).

**Table 2.** Current and pre-existing mental health: asthma vs. no asthma

| **G1-Index Children** |  |  |  |  |  |
| --- | --- | --- | --- | --- | --- |
|  | No Asthma at 28 years<br>N=2532 (86.1%) |  | Asthma at 28 years<br>N=410 (13.9%) |  |  |
|  | Total | Mean (SD) | Total | Mean (SD) | Pvalue† |
| **Pre-existing Mental Health** |  |  |  |  |  |
| WEMWBS-14 Wellbeing score at 24 years | 1959 | 48.99 (8.71) | 313 | 47.54 (9.37) | 0.007 |
| SMFQ-13 Depression score at 26 years | 1979 | 6.63 (6.23) | 313 | 8.09 (6.64) | 1.56E-04 |
| GAD-7 Anxiety score at 22 years | 1615 | 4.50 (4.41) | 245 | 4.87 (4.43) | 0.22 |
| **Mental Health during lockdown** |  |  |  |  |  |
| WEMWBS-14 Wellbeing score at 28 years | 2399 | 44.38 (8.37) | 392 | 42.18 (8.83) | 1.93E-06 |
| SMFQ-13 Depression score at 28 years | 2148 | 6.67 (5.28) | 362 | 8.30 (6.15) | 1.62E-07 |
| GAD-7 Anxiety score at 28 years | 2113 | 6.87 (5.02) | 362 | 8.27 (5.43) | 1.90E-06 |
| **G0 Parents** |  |  |  |  |  |
|  | No Asthma at 59 years<br>N=3337 (89.3%) |  | Asthma at 59 years<br>N=400 (10.7%) |  |  |
|  | Total | Mean (SD) | Total | Mean (SD) | Pvalue† |
| **Pre-existing Mental Health** |  |  |  |  |  |
| EPDS-10 Depression score at 52 years | 2591 | 6.28 (5.27) | 307 | 6.88 (5.33) | 0.06 |
| STAI-20 Anxiety score at 39 years | 2831 | 34.99 (10.18) | 332 | 35.70 (10.56) | 0.23 |
| **Mental Health during lockdown** |  |  |  |  |  |
| WEMWBS-14 Wellbeing score at 59 years | 3087 | 48.11 (8.42) | 364 | 48.09 (8.78) | 0.96 |
| SMFQ-13 Depression score at 59 years | 3076 | 2.89 (3.58) | 364 | 3.61 (4.40) | 4.49E-04 |
| GAD-7 Anxiety score at 59 years | 3138 | 3.36 (4.00) | 371 | 4.20 (4.87) | 2.32E-04 |

† Z-test from logistic regression

Well-being, depression and anxiety levels were all worse in G1 participants with asthma compared to G0 participants with asthma during COVID-19 lockdown (p value <0.001, see **Figures E1, E2, E3**).

In addition to the absolute mental health scores in participants with asthma vs. without asthma, we examined the expected difference in mental health scores during COVID-19 between participants with asthma and without asthma, after adjusting for pre-existing mental health, gender, age, smoking and being overweight. Asthma was associated with a 13% increase in depression score in ALSPAC-G1 (adjusted SCR 1.13 95%CI (1.04,1.22), p=0.005 in ALSPAC-G1) and 15% increase in ALSPAC-G0 (1.15 (1.00,1.31), p=0.05), compared to no asthma. Anxiety scores in people with asthma increased by 14% in ALSPAC-G1 (1.14 (1.04,1.26), p=0.005) and by 16% in ALSPAC-G0 (1.16 (1.02,1.32), p=0.02 in ALSPAC-G0), compared to no asthma. Well-being score in people with asthma decreased by 3% (0.97 (0.95,1.00), p=0.02) in ALSPAC-G1, (NA in ALSPAC-G0), compared to no asthma (**Figure 1** & **Supplementary Tables E2 & E3**). Asthma was associated with a similar increase of anxiety and depression scores during COVID-19 in both generations (Z test p values >0.80. See **Figure 2**, **Table E4**).

**Figure 2.**
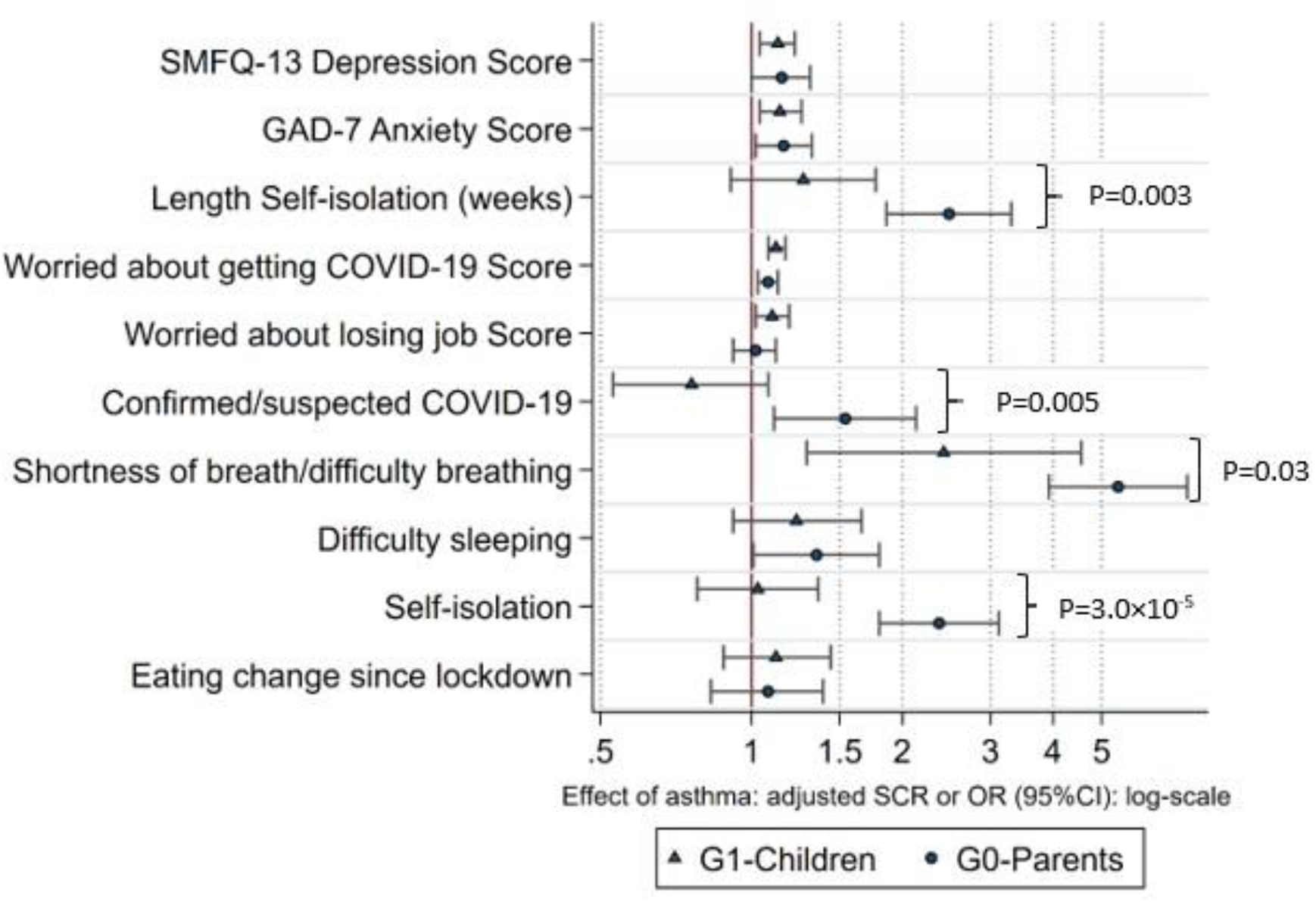
Effect of current asthma on factors specific to lockdown adjusted for age, gender, smoking status, overweight and pre-existing mental health. P-values are from Z-test for 2-sample (2-sided).

### Symptoms, Change in Activities and Worries during lockdown

Since lockdown, ALSPAC-G1 participants with asthma reported more shortness of breath or difficulty in breathing, more difficulty sleeping, longer self-isolation, more eating and sleep changes, more worry about getting COVID-19 and losing their job compared to those without asthma (**Table 3**). The older ALSPAC-G0 participants with asthma also reported more shortness of breath, difficulty sleeping, longer self-isolation, more sleep and eating changes and more worry about getting COVID-19, compared to those without asthma. Additionally, the older G0-ALSPAC participants with asthma reported more confirmed or suspected COVID-19 and more self-isolation, compared to no asthma (**Table 4**).

**Table 3.**
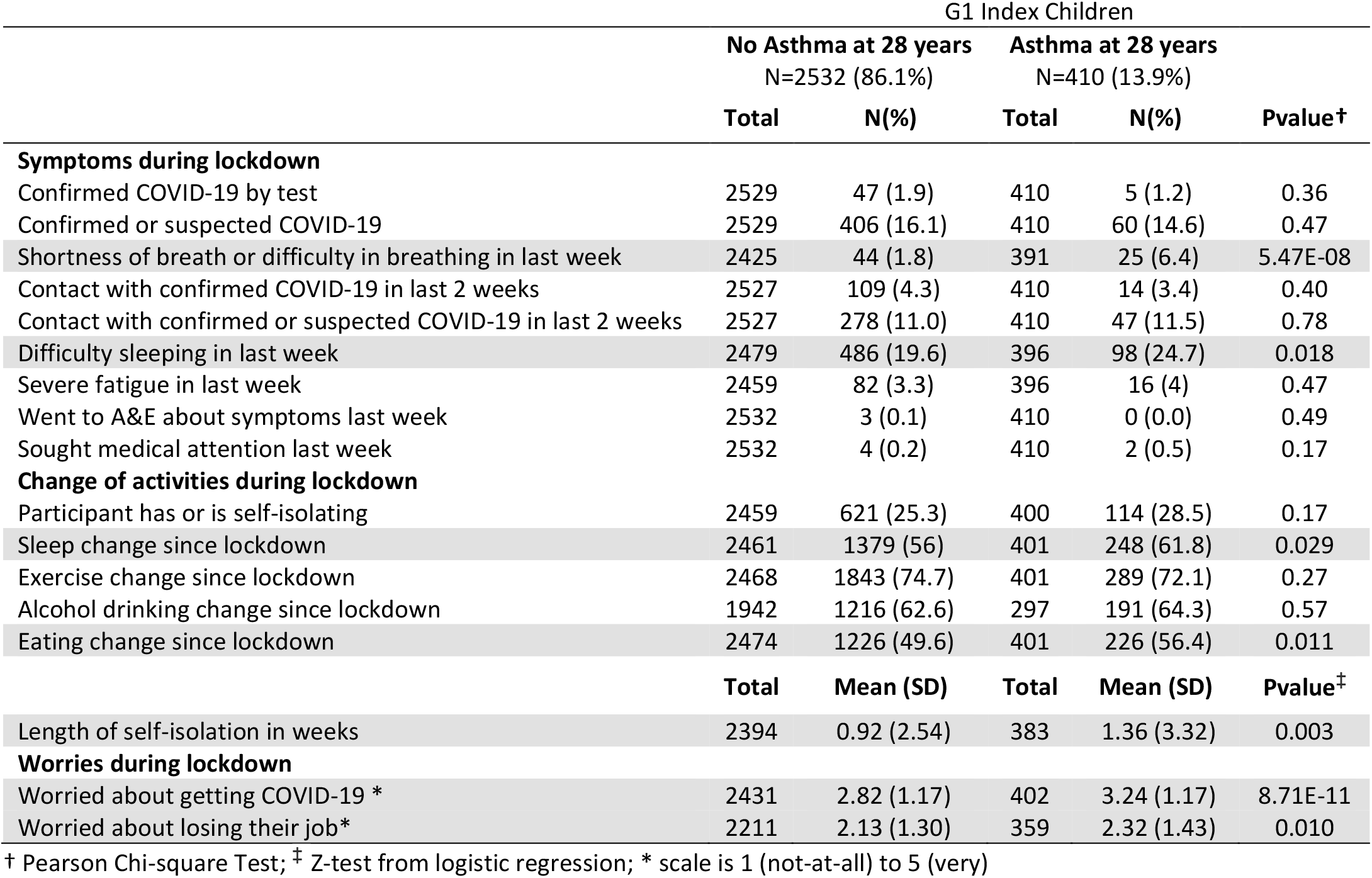
Lockdown characteristics: asthma vs. no asthma in G1-Index Children

**Table 4.**
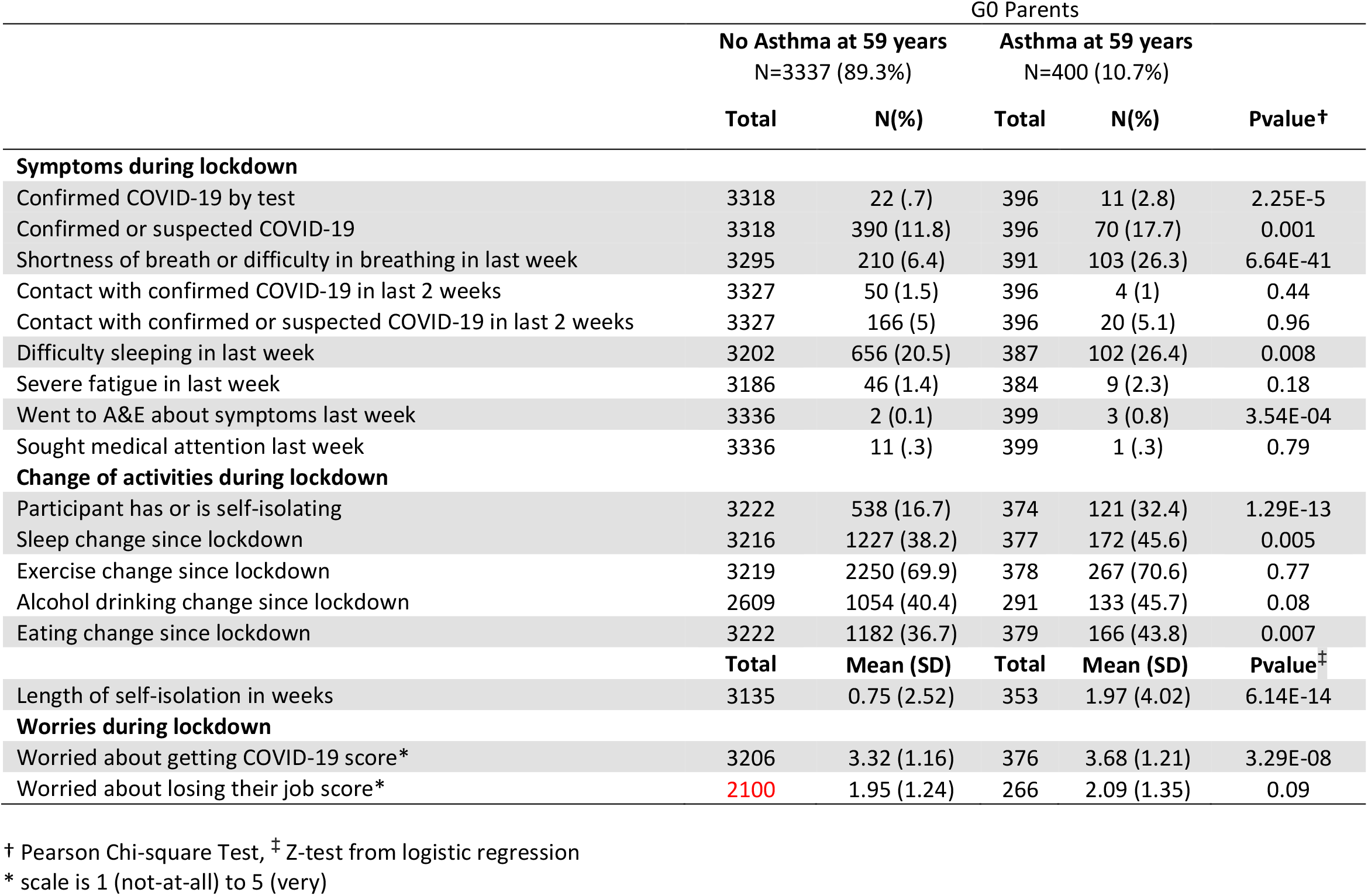
Lockdown characteristics: asthma vs. no asthma in G0 Parents

When comparing the magnitude of the asthma effects in G1 and G0 populations, the older ALSPAC-G0 participants with asthma reported more confirmed or suspected COVID-19 (Z-test p-value G1 vs G0=0.005), more shortness of breath (Z-test p=0.03), more self-isolation (Z-test p=3.0×10^-5^) and longer self-isolation (Z-test p=0.003), compared to the younger ALSPAC-G1 participants with asthma. (**Figure 1** & **Supplementary Tables E2 & E3**).

## Discussion

Surveys suggest that the mental health and wellbeing of populations across the world have been affected by the COVID-19 pandemic [9, 10]. We hypothesised asthma was associated with a greater mental health impact in those with asthma than those without asthma during COVID-19, and that this difference could be related to the psychological effects of the outbreak rather than the physical symptoms of asthma. Our longitudinal study provides evidence that levels of anxiety, mood and wellbeing in the whole ALSPAC population worsened during the pandemic. However, having asthma was associated with a greater decline in mental health, with the younger participants with asthma reporting the worst mental health and wellbeing scores. The data highlights that a range of psychological factors might be mediating this difference, and that these differ by age.

During lockdown, those with asthma in the younger ALSPAC-G1 population had higher levels of depression & anxiety and lower well-being scores than those without asthma. These differences were also evident pre-lockdown for well-being and depression but not anxiety. We identified worse scores for anxiety and depression in the older ALSPAC-G0 participants with asthma, compared to participants without asthma; these differences were not present pre-outbreak with the available scores (different scores to the ones used at the COVID-19 survey). The size of the increase in anxiety and depression scores in asthma compared to those without asthma in the two generations was similar.

The mean WEMWBS-14 score for the younger ALSPAC-G1 participants with asthma dropped from 47.5 to 42.2 across the COVID-19 outbreak, whereas the participants without asthma dropped from 49.0 to 44.4. This decrease in well-being is clinically meaningful for both groups, with mean scores declining to below the cut off for identifying patients at risk of depression in those with asthma [21]. The minimal clinically important difference for the GAD-7 anxiety score has been estimated as 4 [22] and a score of 10+ has a high sensitivity and specificity for the diagnosis of general anxiety disorder [23]. The mean magnitude of increase (post minus pre-COVID-19 score) in the GAD-7 score was 3.4 (mean absolute score during lockdown 8.27) for those with asthma and 2.4 (6.9) for those without asthma in ALSPAC-G1. Therefore, although this does not reach a meaningful clinical difference, with repeated lockdowns it may.

The older ALSPAC-G0 participants with asthma reported more breathlessness and had more confirmed or suspected COVID-19 than the younger ALSPAC-G1 participants with asthma when compared with their peers without asthma. We do not have pre-lockdown measurements of breathlessness, so we cannot say whether the high levels of breathlessness in participants with asthma are pre-existing. If symptoms of respiratory disease were the cause of the anxiety, then we would have expected the older ALSPAC-G0 participants with asthma to be the most anxious group. Whilst we have no direct record of asthma control in our data, a British Lung Foundation survey has estimated that 24.6% of people with asthma triggered by pollution reported an improvement in their symptoms since lockdown [24]. Furthermore, control of asthma in paediatric patients has generally improved since the outbreak [25, 26]. This reduction in asthma symptoms may relate to a decline in pollution levels, improved medication adherence or the interruption of transmission of other viruses that cause exacerbations. This supports our findings and suggest that patients with asthma are not more anxious because of their physical symptoms.

We identified other factors that differed in the population with asthma which might provide more detailed insight into the origins of the increased anxiety. Participants with asthma in the younger ALSPAC-G1 group reported more concerns about catching COVID-19 and worry of losing their jobs when compared with those without asthma. Participants with asthma in the older ALSPAC-G0 group reported increased frequency and length of self-isolation and increased worry of catching COVID-19. On the 16^th^ March 2020 British Prime Minister Boris Johnson announced that vulnerable groups of people will need to shield from social contact for 12 weeks [27]. Such groups at increased risk of severe illness from COVID-19 included those with mild-moderate asthma [28]. This guidance might result in participants with asthma feeling more isolated and having greater concerns for their physical and financial health than those without asthma. The implementation of social isolation measures and the way that they are communicated to the general public may have significantly affected the mental health of patients with asthma more so than those without asthma.

### Strengths & Limitations

The strength of our study lies in the detailed data from the large ALSPAC population. The longitudinal data has provided not just a snapshot of the state of mind of the population during the crisis but has allowed us to assess its development from a pre-pandemic baseline state. The detailed questionnaires also provide us with details about specific concerns during lockdown.

There are limitations to our data. Asthma diagnoses were self-reported, so may over- or under-represent the true asthma population within the group. Whilst this might confound the effect of the physical symptoms of asthma on mental health during the pandemic, it will not affect the psychological outcome of those carrying the label of an asthma diagnosis that is central to our hypothesis. Furthermore, we have evidence of the validity of the self-reported asthma in ALSPAC [29].

The ALSPAC-G0 pre-pandemic assessments of anxiety and depression were made using different assessment tools to those used in the COVID-19 assessment, but we have still managed to assess the mental state of this population over time.

It is difficult to say whether the increase in anxiety and decrease in mental wellbeing in the participants with asthma translates to a significant pathological effect. We do not have data regarding healthcare utilisation of the cohorts pre- and post-lockdown. We are unsure whether people have sought help for their asthma or mental health issues, or whether there has been reduced contact with mental health services for those patients already in the system.

### Future Research

This study highlights higher levels of anxiety and persistently worse markers of depression and mental wellbeing in participants with asthma during the COVID-19 pandemic. This is married with an increased frequency and length of self-isolation and more prominent concerns for physical and financial health. People with asthma are potentially at higher risk of severe COVID-19 infections and it is vital to protect the physical health of this population. Yet there is mixed evidence regarding this risk [30], and the psychological health of this population must not be forgotten. Further investigation is required to clarify the relationship between asthma and COVID-19 infection. It is also important to further clarify the origins of mental health issues in people with asthma. This information should remind the healthcare profession to screen people with asthma for symptoms of anxiety and depression. It will also help to inform government policies which whilst intended to protect the population, are not without negative consequences.

## Conclusion

In conclusion, this study highlights the increased prevalence of anxiety and reduced mental wellbeing in a population with asthma following the start of the COVID-19 pandemic. Clinicians should be made aware of this, and further investigation is required to help inform national policies to try to prevent it.

## Data Availability

The study website contains details of all data available through a fully searchable data dictionary (http://www.bristol.ac.uk/alspac/researchers/our-data/).

## Acknowledgments

We are extremely grateful to all the families who took part in the ALSPAC study, the midwives for their help in recruiting them, and the whole ALSPAC team, which includes interviewers, computer and laboratory technicians, clerical workers, research scientists, volunteers, managers, receptionists and nurses.

## SUPPLEMENTARY INFORMATION

**Appendix 1**. ALSPAC description

**Appendix 2**. Baseline Mental Health Measures

**Appendix 3**. Supplementary tables

**Appendix Table E1**. Pre-COVID-19 asthma in G1-Index children and G0 Parents and lung function in G1-Index children
**Appendix Table E2**. Effect of current asthma on outcomes specific to lockdown adjusted for age, gender, smoking, overweight and pre-existing mental health in G1-Index Children (as shown in Figure 1)
**Appendix Table E3**. Effect of current asthma on outcomes specific to lockdown adjusted for age, gender, smoking, overweight and pre-existing mental health in G0-parents (as shown in Figure 1)
**Table E4**. Comparison of asthma-effects in ALSPAC-G0 vs. ALSPAC-G1 (as shown in Figure 1)

**Appendix 4**. Supplementary Figures

**Figure E1**. WEMWBS-17 wellbeing during lockdown in G0-ALSPAC children with asthma vs G1-ALSPAC parents with asthma
**Figure E2**. SMFQ-13 Depression score during lockdown in G0-ALSPAC children with asthma vs G1-ALSPAC parents with asthma
**Figure E3**. GAD-7 Anxiety score during lockdown in G0-ALSPAC children with asthma vs G1-ALSPAC parents with asthma

**Appendix 5**. References

## Appendix 1. ALSPAC Description

ALSPAC recruited 14,541 pregnant women resident in Avon, UK with expected dates of delivery 1st April 1991 to 31st December 1992. 14,541 is the initial number of pregnancies for which the mother enrolled in the ALSPAC study and had either returned at least one questionnaire or attended a “Children in Focus” clinic by 19/07/99. Of these initial pregnancies, there was a total of 14,676 fetuses, resulting in 14,062 live births and 13,988 children who were alive at 1 year of age.

When the oldest children were approximately 7 years of age, an attempt was made to bolster the initial sample with eligible cases who had failed to join the study originally. As a result, when considering variables collected from the age of seven onwards (and potentially abstracted from obstetric notes) there are data available for more than the 14,541 pregnancies mentioned above.

The number of new pregnancies not in the initial sample (known as Phase I enrolment) that are currently represented on the built files and reflecting enrolment status at the age of 18 is 706 (452 and 254 recruited during Phases II and III respectively), resulting in an additional 713 children being enrolled. The phases of enrolment are described in more detail in the cohort profile paper: <http://ije.oxfordjournals.org/content/early/2012/04/14/ije.dys064.full.pdf+html>.

The total sample size for analyses using any data collected after the age of seven is therefore 15,247 pregnancies, resulting in 15,458 fetuses. Of this total sample of 15,458 fetuses, 14,775 were live births and 14,701 were alive at 1 year of age.

A 10% sample of the ALSPAC cohort, known as the Children in Focus (CiF) group, attended clinics at the University of Bristol at various time intervals between 4 to 61 months of age. The CiF group were chosen at random from the last 6 months of ALSPAC births (1432 families attended at least one clinic). Excluded were those mothers who had moved out of the area or were lost to follow-up, and those partaking in another study of infant development in Avon.

The study website contains details of all data available through a fully searchable data dictionary (http://www.bristol.ac.uk/alspac/researchers/our-data/). Ethical approval for the study was obtained from the ALSPAC Ethics and Law Committee and the Local Research Ethics Committees. Informed consent for the use of data collected via questionnaires and clinics was obtained from participants following the recommendations of the ALSPAC Ethics and Law Committee at the time.

Informed consent for the use of data collected via questionnaires and clinics was obtained from participants following the recommendations of the ALSPAC Ethics and Law Committee at the time.

Study data were collected and managed using REDCap electronic data capture tools [1, 2] hosted at University of Bristol.

## Appendix 2. Baseline Mental Health Measures

Depressive symptoms: Baseline depressive symptoms were measured using the Edinburgh Postnatal Depression Scale or EPDS, [3] between 2011-2013. The EPDS consists of 10 items probing depressive symptoms in the last two weeks with scores ranging between 0-30 and higher scores indicating greater depression. Scores ≥ 12 have been validated against probable major depression.[4]

Anxiety symptoms: Baseline anxiety symptoms were measured using the Spielberger State Trait Anxiety Inventory or STAI, [5] between 1999-2001. We used the trait scale of the STAI, which consists of 20 items with scores ranging between 20-80 with higher scores indicating greater trait anxiety. Scores ≥ 46 have been validated to predict a moderate anxiety disorder. [5]

## Appendix 3. Supplementary Tables

**Table E1.**
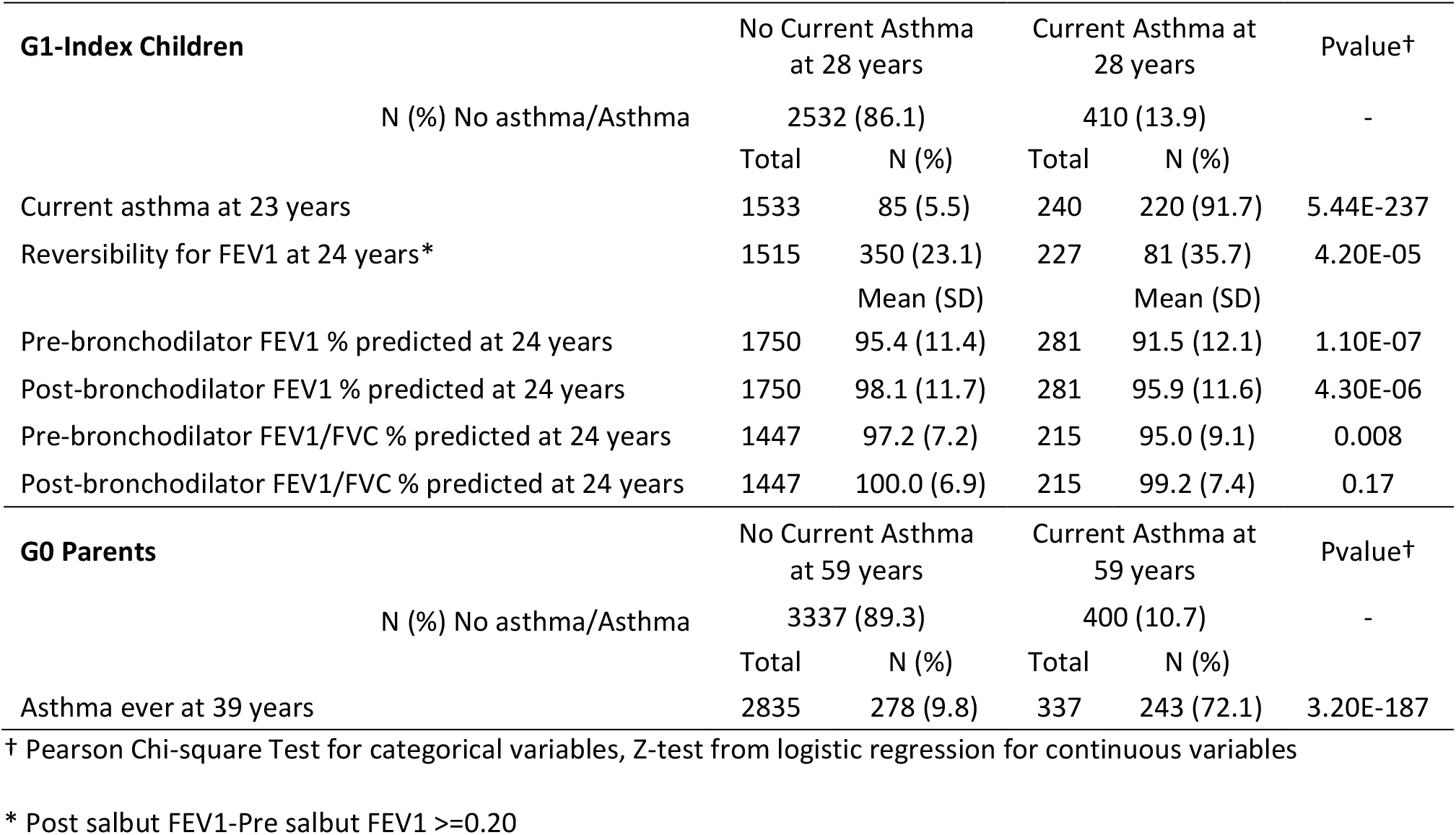
Pre-COVID-19 asthma in G1-Index children and G0 Parents and lung function in G1-Index children

**Table E2.** Effect of current asthma on outcomes specific to lockdown adjusted for age, gender, smoking, overweight and pre-existing mental health in G1-Index Children (as shown in Figure 1)

| Effect of Current Asthma at 27 years on Lockdown in G1-Index Children |  |  |  |  |  |  |  |  |
| --- | --- | --- | --- | --- | --- | --- | --- | --- |
|  | R<sup>2</sup> | Crude Model-restricted | Pvalue | N | R<sup>2</sup> | Adjusted Model* | Pvalue | N |
| **Count Outcomes** |  | SCR (95% CI) |  |  |  | SCR (95% CI) |  |  |
| WEMWBS-14 Wellbeing score (14-70) | 0.001 | 0.96 (0.93,0.98) | 0.001 | 2153 | 0.05 | 0.97 (0.95,1.00) | 0.016 | 2153 |
| SMFQ-13 Depression score (0-26) | 0.007 | 1.26 (1.14,1.38) | 3.03E-06 | 1963 | 0.16 | 1.13 (1.04,1.22) | 0.005 | 1963 |
| GAD-7 Anxiety score (0-21) | 0.003 | 1.17 (1.06,1.29) | 0.002 | 1557 | 0.08 | 1.14 (1.04,1.26) | 0.005 | 1557 |
| Length of self-isolation in weeks (0-13) | 0.002 | 1.32 (0.95,1.84) | 0.09 | 2144 | 0.02 | 1.27 (0.91,1.77) | 0.15 | 2144 |
| Worried about getting COVID-19 score (1-5) | 0.002 | 1.14 (1.09,1.19) | 6.27E-09 | 2186 | 0.01 | 1.12 (1.08,1.17) | 7.55E-08 | 2186 |
| Worried about losing their job score (1-5) | 0.001 | 1.12 (1.03,1.21) | 0.005 | 1980 | 0.005 | 1.10 (1.02,1.19) | 0.016 | 1980 |
| **Binary Outcomes** |  | OR (95%CI) |  |  |  | OR (95%CI) |  |  |
| Confirmed or suspected COVID19 | 0.00 | 0.76 (0.54,1.08) | 0.13 | 2218 | 0.00 | 0.76 (0.53,1.08) | 0.13 | 2218 |
| Shortness of breath or difficulty in breathing | 0.02 | 2.81 (1.51,5.21) | 0.001 | 2136 | 0.05 | 2.42 (1.29,4.55) | 0.01 | 2136 |
| Difficulty sleeping | 0.00 | 1.34 (1.01,1.79) | 0.046 | 2170 | 0.03 | 1.23 (0.92,1.66) | 0.17 | 2170 |
| Participant has or is self-isolating | 0.00 | 1.08 (0.81,1.42) | 0.61 | 2202 | 0.01 | 1.03 (0.78,1.36) | 0.84 | 2202 |
| Eating change since lockdown | 0.00 | 1.20 (0.94,1.54) | 0.13 | 2210 | 0.02 | 1.12 (0.88,1.44) | 0.36 | 2210 |

† adjusted by pre-existing mental health, overweight, smoking, gender and age; \* scale is 1 (not-at-all) to 5 (very); ‡ WEMWBS score not available for G0-ALSPACSCR=symptom count ratios from Poisson regression models; OR=odds ratios from logistic regression models

**Table E3.** Effect of current asthma on outcomes specific to lockdown adjusted for age, gender, smoking, overweight and pre-existing mental health in G0-parents (as shown in Figure 1)

| Effect of Current Asthma at 59 years on Lockdown in G0-Parents |  |  |  |  |  |  |  |  |
| --- | --- | --- | --- | --- | --- | --- | --- | --- |
|  | R<sup>2</sup> | Crude Model-restricted | Pvalue | N | R<sup>2</sup> | Adjusted Model* | Pvalue | N |
| **Count Outcomes** |  | SCR (95%CI) |  |  |  | SCR (95%CI) |  |  |
| SMFQ-13 Depression score (0-26) | 0.002 | 1.23 (1.06,1.43) | 0.007 | 2652 | 0.14 | 1.15 (1.00,1.31) | 0.05 | 2652 |
| GAD-7 Anxiety score (0-21) | 0.003 | 1.26 (1.10,1.45) | 9.97E-04 | 2940 | 0.11 | 1.16 (1.02,1.32) | 0.02 | 2940 |
| Length of self-isolation in weeks (0-13) | 0.024 | 2.52 (1.89,3.36) | 2.87E-10 | 2689 | 0.05 | 2.48 (1.86,3.30) | 5.8E-10 | 2689 |
| Worried about getting COVID-19 score (1-5) | 0.001 | 1.09 (1.04,1.14) | 9.44E-05 | 2758 | 0.004 | 1.08 (1.03,1.13) | 4.1E-04 | 2758 |
| Worried about losing their job score (1-5) | 0.000 | 1.04 (0.94,1.15) | 0.41 | 1784 | 0.01 | 1.02 (0.92,1.12) | 0.73 | 1784 |
| **Binary Outcomes** |  | OR (95%CI) |  |  |  | OR (95%CI) |  |  |
| Confirmed or suspected COVID19 | 0.00 | 1.63 (1.18,2.25) | 0.003 | 2770 | 0.01 | 1.54 (1.11,2.13) | 0.01 | 2770 |
| Shortness of breath or difficulty in breathing | 0.07 | 5.75 (4.21,7.84) | 3.01E-28 | 2748 | 0.10 | 5.39 (3.92,7.41) | 3.4E-25 | 2748 |
| Difficulty sleeping | 0.00 | 1.45 (1.09,1.93) | 0.01 | 2685 | 0.04 | 1.35 (1.01,1.80) | 0.04 | 2685 |
| Participant has or is self-isolating | 0.01 | 2.39 (1.82,3.13) | 3.92E-10 | 2769 | 0.02 | 2.37 (1.80,3.12) | 8.5E-10 | 2769 |
| Eating change since lockdown | 0.00 | 1.20 (0.94,1.54) | 0.150 | 2771 | 0.04 | 1.08 (0.83,1.39) | 0.57 | 2771 |

† adjusted by pre-existing mental health, overweight, smoking, gender and age. In the parents, WEMWBS-14 Wellbeing score is not available at a previous timepoint. We used previous STAI score at 39 years to adjust current GAD-7 anxiety at 59 years. For all other variables we used previous EPDS score for depression at 52 years to adjust for pre-existing mental health. \* scale is 1 (not-at-all) to 5 (very)
SCR= symptom count ratios from Poisson regression models; OR=odds ratios from logistic regression models

**Table E4.** Comparison of asthma-effects in ALSPAC-G0 vs. ALSPAC-G1 (as shown in Figure 1)

|  | G0-Parents |  | G1-Children |  | Coef_G1-Coef_G0 | SE_G1-SE_G0 |  |  |
| --- | --- | --- | --- | --- | --- | --- | --- | --- |
| **Count Outcomes** | **Coef1** | **SE1** | **Coef1** | **SE1** | **Coef_diff** | **SE_diff** | **Z=Coeff_diff/SE_diff** | **Z test**<br>**P-value** |
| SMFQ-13 Depression score (0-26) | 0.14 | 0.07 | 0.12 | 0.04 | -0.02 | 0.08 | -0.25 | 0.80 |
| GAD-7 Anxiety score (0-21) | 0.15 | 0.07 | 0.13 | 0.05 | -0.02 | 0.09 | -0.23 | 0.82 |
| Length of self-isolation in weeks (0-13) | 0.91 | 0.15 | 0.24 | 0.17 | -0.67 | 0.23 | -2.96 | 0.003 |
| Worried about getting COVID-19 score (1-5) | 0.08 | 0.02 | 0.12 | 0.02 | 0.04 | 0.03 | 1.41 | 0.16 |
| Worried about losing their job score (1-5) | 0.02 | 0.05 | 0.10 | 0.04 | 0.08 | 0.06 | 1.25 | 0.21 |
| **Binary Outcomes** | **Coef2** | **SE2** | **Coef2** | **SE2** | **Coef_diff** | **SE_diff** | **Z=Coeff_diff/SE_diff** | **Z test**<br>**P-value** |
| Confirmed or suspected COVID19 | 0.43 | 0.17 | -0.27 | 0.18 | -0.70 | 0.25 | -2.83 | 0.005 |
| Shortness of breath or difficulty in breathing | 1.68 | 0.16 | 0.88 | 0.32 | -0.80 | 0.36 | -2.24 | 0.025 |
| Difficulty sleeping | 0.3 | 0.15 | 0.21 | 0.15 | -0.09 | 0.21 | -0.42 | 0.67 |
| Participant has or is self-isolating | 0.86 | 0.14 | 0.03 | 0.14 | -0.83 | 0.20 | -4.19 | 3.00E-05 |
| Eating change since lockdown | 0.07 | 0.13 | 0.11 | 0.13 | 0.04 | 0.18 | 0.22 | 0.83 |
| Felt miserable or unhappy in past 2 weeks | 0.14 | 0.13 | 0.18 | 0.15 | 0.04 | 0.20 | 0.20 | 0.84 |
| Felt lonely in past 2 weeks | 0.32 | 0.14 | 0.11 | 0.13 | -0.21 | 0.19 | -1.10 | 0.27 |

Coef1, SE1 are the logarithmic form of the SRC and corresponding standard error; Coef2, SE2 are the logarithmic form of the OR and corresponding standard error

## Appendix 4. Supplementary figures

**Figure E1.**
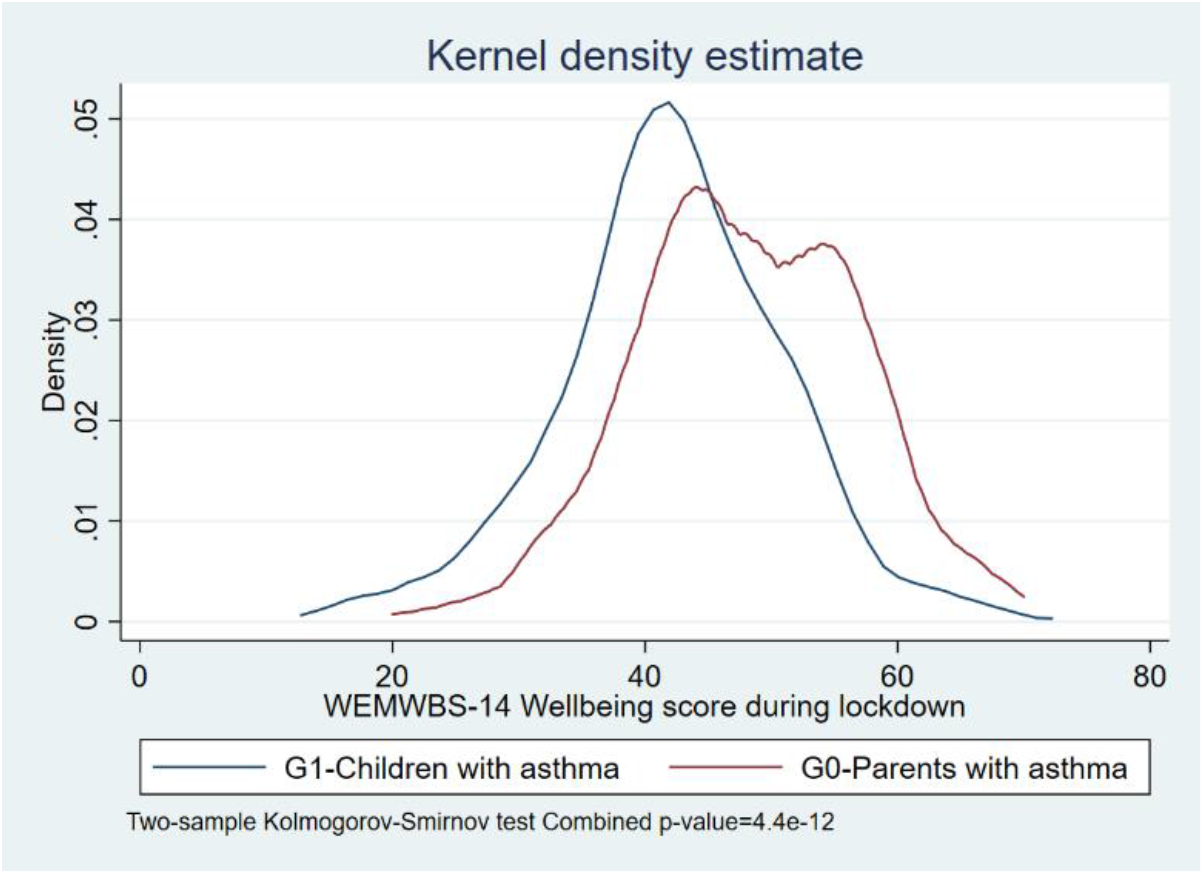
WEMWBS-14 Wellbeing score during lockdown in G0-ALSPAC children with asthma vs G1-ALSPAC parents with asthma

**Figure E2.**
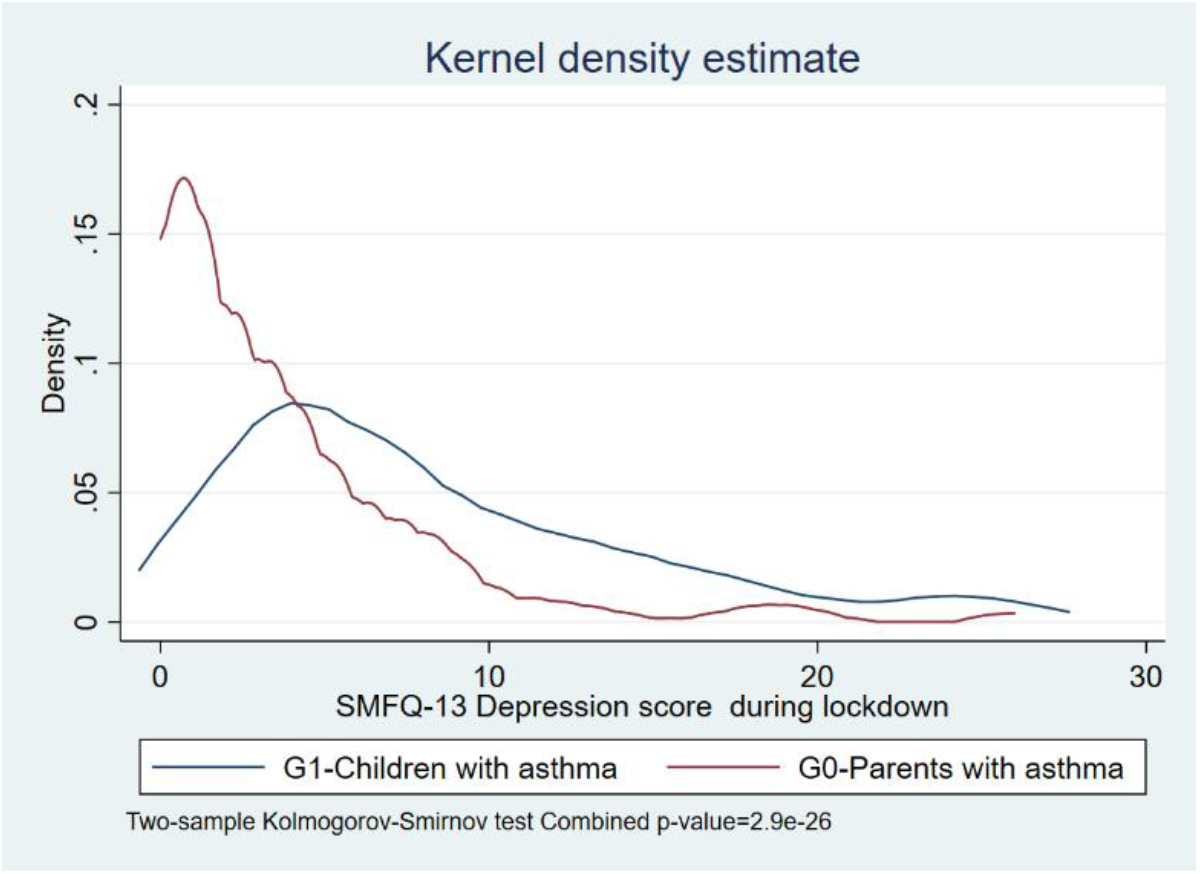
SMFQ-13 Depression score during lockdown in G0-ALSPAC children with asthma vs G1-ALSPAC parents with asthma

**Figure E3.**
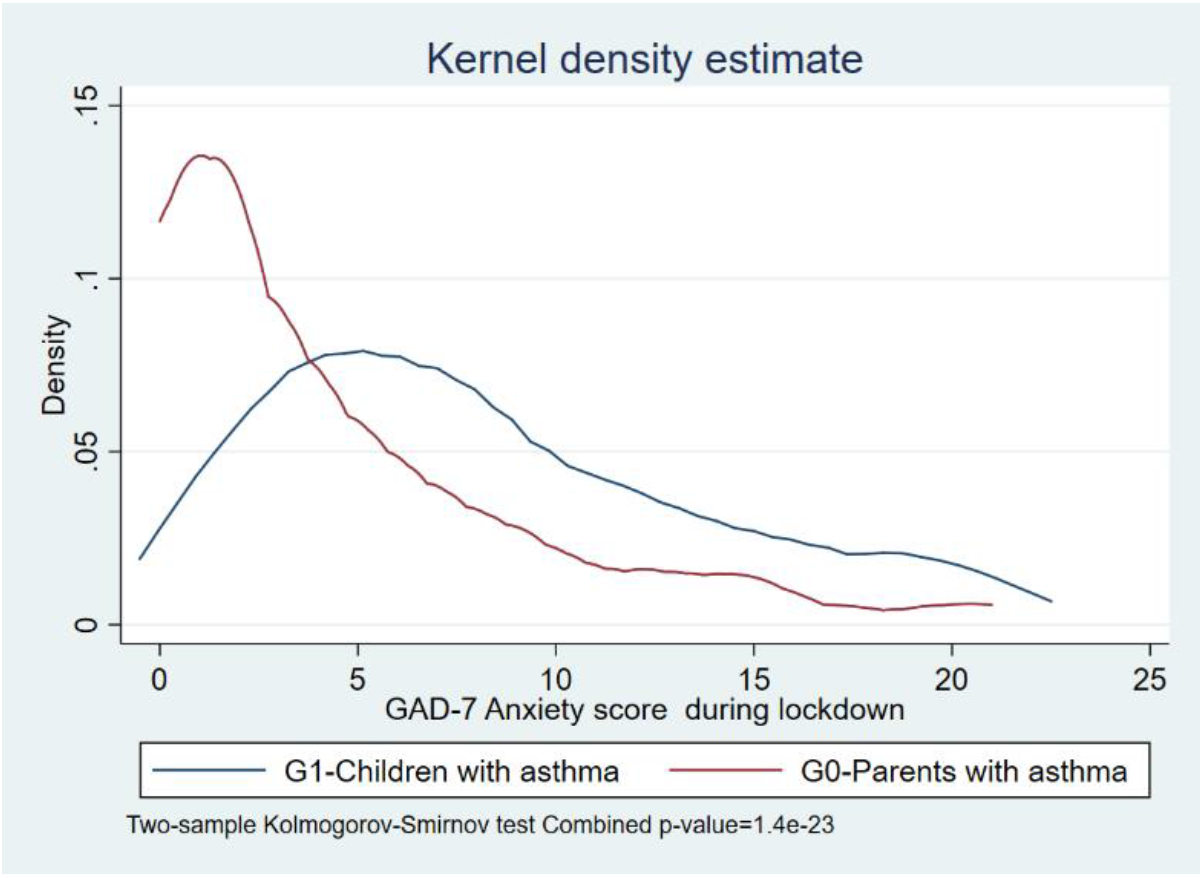
GAD-7 Anxiety score during lockdown in G0-ALSPAC children with asthma vs G1-ALSPAC parents with asthma

## Notes

Funding: This work was supported by the Medical Research Council and the University of Bristol (MC_UU_00011/1). MRC CARP Fellowship. The UK Medical Research Council and Wellcome (Grant ref: 217065/Z/19/Z) and the University of Bristol provide core support for ALSPAC. This publication is the work of the authors and Raquel Granell will serve as guarantors for the contents of this paper. A comprehensive list of grants funding is available on the ALSPAC website (http://www.bristol.ac.uk/alspac/external/documents/grant-acknowledgements.pdf.

### Competing Interest Statement

The authors have declared no competing interest.

### Author Declarations

Ethical approval for the study was obtained from the ALSPAC Ethics and Law Committee and the Local Research Ethics Committees. Informed consent for the use of data collected via questionnaires and clinics was obtained from participants following the recommendations of the ALSPAC Ethics and Law Committee at the time. Informed consent for the use of data collected via questionnaires and clinics was obtained from participants following the recommendations of the ALSPAC Ethics and Law Committee at the time.

